# High accuracy indicators of androgen suppression therapy failure for prostate cancer – a modeling study

**DOI:** 10.1101/2022.06.24.22276874

**Authors:** William Meade, Allison Weber, Tin Phan, Emily Hampston, Laura Figueroa Resa, John Nagy, Yang Kuang

**Author notes:** These authors contributed equally (William Meade, Allison Weber, Tin Phan).

## Abstract

Prostate Cancer is a serious public health concern in the United States. The primary obstacle to effective long-term management for prostate cancer patients is the eventual development of treatment resistance. Due to the uniquely chaotic nature of the neoplastic genome, it is difficult to determine the evolution of tumor composition over the course of treatment. Hence, a drug is often applied continuously past the point of effectiveness, thereby losing any potential treatment combination with that drug permanently to resistance. If a clinician is aware of the timing of resistance to a particular drug, then they may have a crucial opportunity to adjust the treatment to retain the drug usefulness in potential treatment combination or strategy. In this study, we investigate new methods of predicting treatment failure due to treatment resistance using a novel mechanistic model built on an evolutionary interpretation of Droop cell quota theory. We analyze our proposed methods using patient PSA and androgen data from a clinical trial of intermittent treatment with androgen deprivation therapy. Our results produce two indicators of treatment failure. The first indicator is calculated using our mathematical model with a predictive accuracy of 87.3% (sensitivity: 96.1%, specificity: 65%). The second indicator is calculated directly from serum androgen and PSA data with a predictive accuracy of 88.7% (sensitivity: 90.2%, specificity: 85%). The high sensitivity of the first indicator and the high specificity of the second indicator means they can complement one another in clinical settings. Our results demonstrate the potential and feasibility of using evolutionary tumor dynamics models in combination with the appropriate data to aid in the adaptive management of prostate cancer.

## Introduction

Prostate cancer is the most prevalent cancer among men in the US, and therefore is a significant public health concern (1). Treatment for prostate cancer has advanced considerably over the past several decades (20), but treatment resistance remains a significant threat to every existing therapy. The high degree of heterogeneity in prostate cancer means that even treatments with a promising initial response can fail when the neoplasm inevitably evolves to become treatment resistant (21,22). Furthermore, applying any therapy to the point of failure only serves to further fortify the existing resistance. If it were possible to identify an incipient resistance then a clinician would have an opportunity to change treatment strategy to potentially reach a more favorable outcome of the overall regimen (15,23,24).

By nature, cancer is often highly genomically unstable. As a result the constituent cells of any neoplasm are endlessly differentiating (2–4). Therefore, tumors tend not to be homogeneous collections of genetically identical cancerous cells (4). New genomic variations arise continually and rapidly, and some will inevitably confer traits that allow them to evade a previously effective therapy (4,48). In the presence of resistant cancer cells, treatment would select for the dominance of the resistant trait. Once the susceptible cells are eliminated, the tumor becomes permanently and irrevocably resistant to that treatment. This clonal model of resistance explains why it is undesirable to continue any therapy to the point of failure (4,5).

There is evidence that treatment resistance comes at a fitness cost, which means resistant phenotypes are unlikely to become dominant in a treatment-free environment (25). Therefore, it has been suggested that one could exploit intracellular competition by adjusting the timing and intensity of treatment to effectively reduce the development of treatment resistant phenotypes and thereby manage the progression of the tumor. This is the central idea of adaptive therapy, deeply rooted in ecological theory, but remains difficult to execute in practice (23-25).

Pretreatment, prostate cancers are androgen-dependent. Androgens diffuse into prostate cells and bind to intracellular androgen receptors, which then activate proliferation and survival pathways in both healthy and cancerous prostate cells. For this reason, androgen deprivation therapy (ADT) is the standard of treatment for advanced or metastatic prostate cancer (26). The therapy uses an agonist and(or) antagonist of luteinizing hormone-releasing factor and antiandrogen drugs to eliminate primary androgen production in the testes, respectively. ADT is initially effective at stopping and reversing the growth of the tumor, but treatment resistance inevitably arises (1,6,13).

The standard application of ADT involves continuously applying the treatment at maximum dosage in order to eradicate the tumor quickly (continuous androgen suppression, or CAS). However, due to the adverse side effects of ADT and the imminent development of treatment resistance, intermittent androgen suppression therapy (IAS) was theorized to be the better alternative. IAS is a rigid form of adaptive therapy, where ADT is applied at maximum dosage on either on- and off-intervals with fixed duration or based on the growth of tumor (7). IAS has several advantages over CAS. Most notably, the off-treatment periods in IAS give patients a break from the adverse side-effects of ADT, hence improving the overall quality of life for patients (27). There was concern regarding the comparative effectiveness of IAS; however, meta-analyses show no statistical difference in time to remission between IAS and CAS (28).

Androgen also triggers secretion of prostate-specific antigen (PSA), a protein normally found in seminal fluid. Usually, PSA is contained within the cytoplasm of prostatic acinar cells and ductal epithelium. However, when the prostate becomes cancerous, PSA can leak into the bloodstream via disruption in the epithelial wall. PSA then can be detected in the serum, where a high level of PSA is a strong indicator of prostate cancer presence and growth. Hence, clinicians can monitor prostate cancer progression using longitudinal measurements of PSA. The correlation between PSA levels and tumor volume is imperfect and can vary over time due to phenotypical and physiological changes in the tumor (29). Furthermore, PSA is not cancer specific. However, since PSA measurements can be taken frequently at low cost, it remains a valuable tool that can be used to gain invaluable insights into the dynamics of the tumor (7,30).

The development of dynamical models for prostate cancer dates back almost two decades (31). Since then, there has been an array of models developed to study different aspects of prostate cancer and its treatments (5,8-13,17,25,32-43). Most of these results have been reviewed and synthesized previously (15,44,45).

In this study, we use a mathematical model to track the clonal evolution of prostate cancer cells. Using longitudinal measurements of androgen and PSA from a clinical trial for IAS, we demonstrate the potentials of two new methods for predicting an imminent treatment failure due to the growing dominance of resistant cellular strains (7). The first method is developed based on the mathematical model, while the second is model-free and based on the underlying theoretical implication of the first method. For the purpose of this work, we classify all prostate cancer cell types into two broad categories: those susceptible and those resistant to ADT. These two indicators of treatment resistance, or biomarkers, each has its own advantages, but both have the potential to be useful in clinical settings.

## Materials and Methods

Our primary investigative tool is a mathematical model based on the Droop cell quota framework and multi-species competition theory (46). The model is presented in detail in the quick-guide box. In summary, the model represents a system wherein androgen is produced and secreted into the blood, before diffusing into the intracellular spaces of cancerous cells in the prostate. The resulting cell quota of androgen, *Q(t)*, is representative of the bound androgen receptors which drive the proliferation and apoptosis of prostate cancer cells. In order to proliferate, cancer cells require a certain amount of bound androgen receptors. This minimum amount of bound receptors is called the minimum cell quota, or *q*, within the Droop framework. For example, if a cell lacks a sufficient number of bound receptors to support proliferation (*Q*(*t*) ≤ *q*), then the proliferation term becomes 0. The Droop functions have been used extensively to model prostate cancer (8–13).

As prostate cancer cells proliferate, they produce PSA. Previous studies considered the PSA production rate to be linearly dependent on the current amount of bound androgen receptors. To be more biologically realistic, we assume that PSA production rate is proportional to the proliferation rate of cancer cells. That is, if cancer cells do not proliferate, then they do not produce PSA. We test the qualitative behaviors of the two assumptions to show that our proposed alternative PSA production rate better captures the qualitative behaviors of PSA dynamics (see model formulation in supplementary material). These considerations for our modeling framework is highlighted in Figure 1.

**Figure 1:**
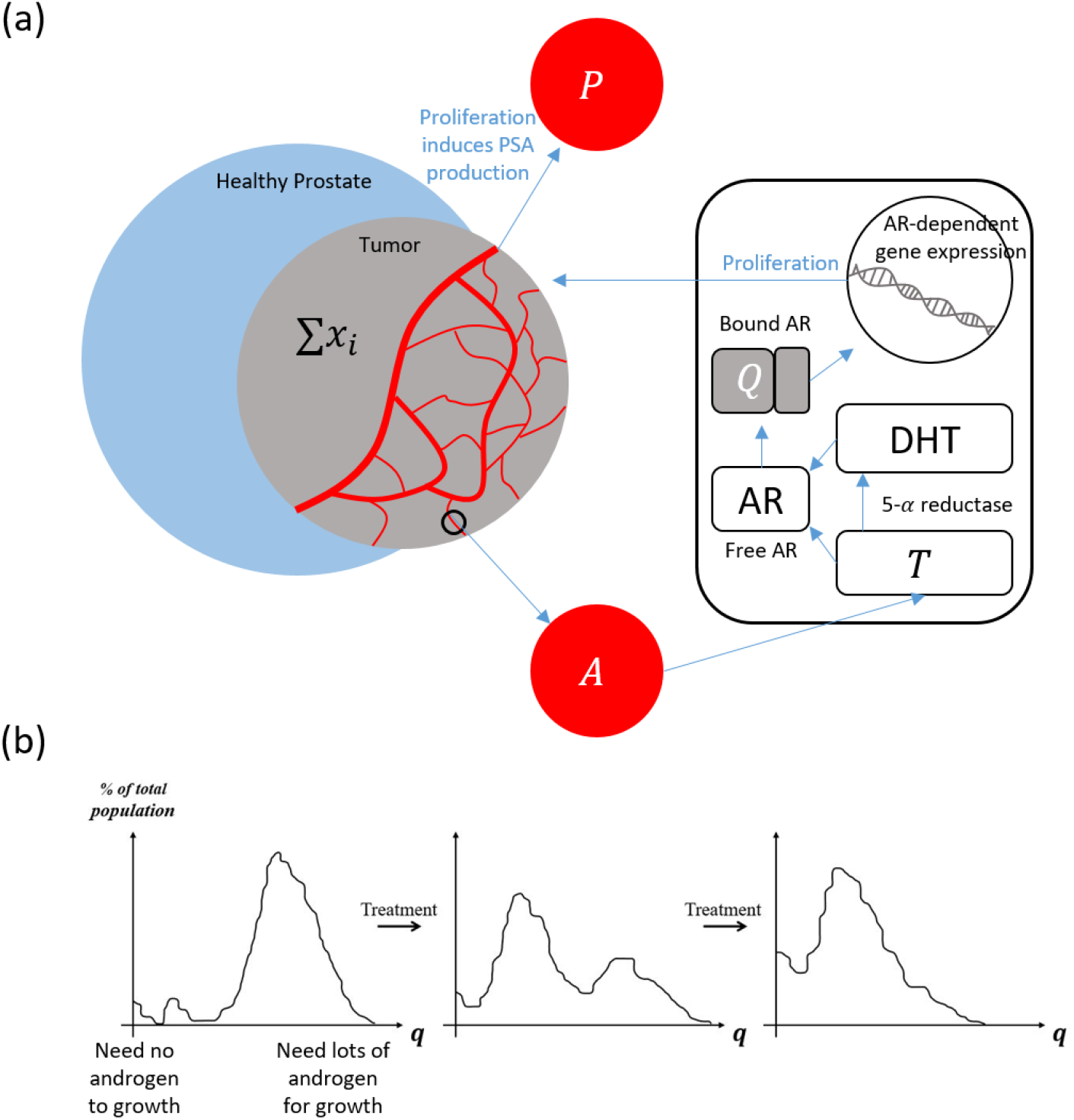
Schematics of model foundation and evolutionary framework. (a) Androgens (testosterones) enter the cancer cells. Some are converted to the potent dihydrotestosterone (DHT) with the help of 5-*α* reductase. Both then bind to the androgen receptors (AR). The bound androgen receptors send proliferative signals for cancer to grow and produce PSA (P). PSA is then leaked to the blood stream. (b) The distribution of the minimum cell quota (q) prior to treatment is skewed to higher values for q. This means most cancer cells are initially sensitive to treatment. During each treatment, this evolutionary landscape shifts toward a lower average q, meaning an increasing number of cells become less dependent on exogenous androgen.

## Model Quick-Guide Box

This model is built on previous work by Kuang et al. (8-13). A schematics is provided in Figure 1.

The total volumes of cancer cells susceptible to treatment (Castration Susceptible or CS) and resistant to treatment (Castration Resistant or CR) are represented by *x_1_* and *x_2_*, respectively. Hence, together 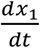 and 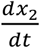 capture the rate of change of the total cancer population as it undergoes intermittent androgen suppression therapy (IAS). Our model of this dynamic takes the following form:

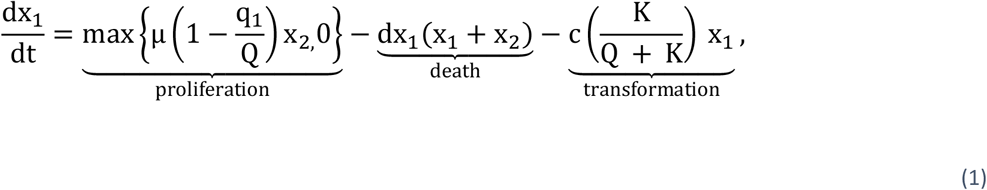

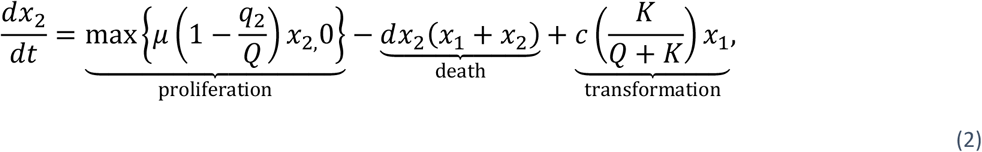

The maximum functions represent androgen-dependent proliferation. In the presence of sufficient androgen (i.e., *Q* > *q*_*i*_), proliferation and PSA production rates are positive. Otherwise, when androgen is below the minimum cell quota level, proliferation and PSA secretion rates go to 0 in our model formulation. Additionally, androgen also affects the transformation rate based on a standard saturation.

The density-dependent death term represents the competition between and among the *x*_1_ and *x*_2_ populations for other resources, including glucose and oxygen. If evolutionary trade-off is to be considered, we may select different competition rates *d*′*s* to reflect the cost of gaining resistance (17,25,32). However, for simplification, the current model assumes that even without treatment, the tumor is guaranteed to become treatment-resistant given a sufficiently long time because we purposefully neglect cost of resistance and the variety of potential subclones in order to simplify the model. This is justified because modern theory of treatment, “hit hard, hit fast,” essentially destroys susceptible clones entirely (44). Our goal here is to investigate treatment resistance under current clinical practice. A more complex model would be required to explore adaptive therapy.

A crucial component to our modeling framework is the pair of minimum cell quota parameters *q*_1_ and *q*_2_ that define the threshold amounts of bound androgen receptor required for proliferation of the 2 subclones. While we elect to have only two subpopulations, the classification is based on a population-average, so the resistance level of *x*_2_ may change over time to account for evolutionary factors. In particular, the resistant population *x*_2_ is expected to decrease its average dependence on androgen as the treatment goes on. This means we expect to see a diminishing *q*_*2*_ as we calibrate its value over the course of treatment see Figure 1.b. This means *x*_2_ represents the currently dominant resistant clone.

Under selective pressure of the treatment, cells may adapt to increase their survivability (i.e., mutations, genomic or epigenetic changes). The transformation term represents the rate at which cells adapt and become more independent of androgen over time. Previous modeling studies have shown that it is not necessary to include a transformation term from resistant to sensitive phenotypes (10). The parameter *K* determines how sensitive this transformation rate is to the level of bound androgen receptors.

The free androgen and bound androgen receptors are represented by *A*(*t*) and *Q*(*t*), respectively. We model their dynamics as follows:

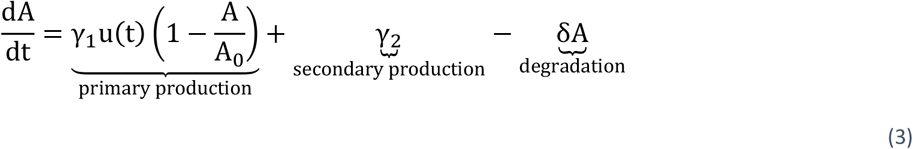

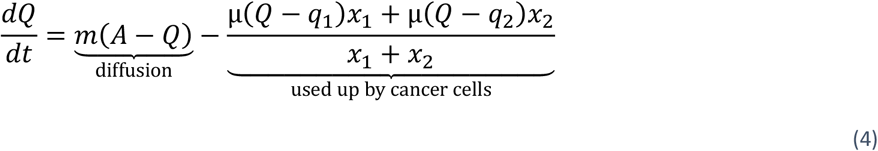

Production of free androgen follows a negative feedback loop with maximum rate parameter γ_1_ and homeostasis androgen level *A*_0_. Testicular production of androgen (primary production) is intermittently suppressed by administration of ADT, which is represented by the Heaviside function *u*(*t*). The rate of adrenal androgen production γ_2_ is constant and fixed at a small percentage of the primary production γ_1_. Serum androgen concentration degrades at constant-per-capita rate δ.

Free androgen diffuses into cells and binds to androgen receptor at a maximum rate *m*. We assume that androgen receptor binding happens instantaneously when androgen enters the cell. The term for the amount of androgen used up by cancer for growth accounts for changes to intracellular androgen concentration due to proliferation, which is derived from conservation laws (10).

In general, existing prostate cancer models are built on the assumption that the rate of PSA production is a linear function of the amount of tumor cells (15). However, PSA production is intrinsically linked to cancer proliferation via the same transcription factor (bound androgen receptors), see Figure 1-a. Thus, we reflect this observation by formulating PSA production rate as a function of cellular activity in our model. In addition, we add a baseline production of PSA due to healthy prostate cells. These assumptions lead to the following equation for PSA dynamics:

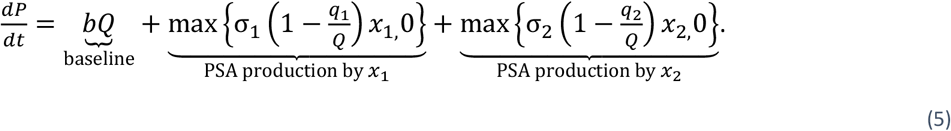

One desirable consequence of connecting the cellular proliferation function to the PSA production rate is the explicit connection between minimum cell quotas, *q*_1_ and *q*_2_, and the level of PSA. As the neoplasm becomes increasingly indifferent to environmental androgen, resistant cancer cells should secrete PSA more freely even during active ADT. We hypothesize that in our model the dynamics of PSA will be sensitive to changes in the *q*_*1*_ and *q*_*2*_ parameters. Additionally, the model should reflect the divergence between PSA and androgen levels observed in later cycles of resistant patients.

Table 1 contains a summary of model parameters and ranges. Additional information on the model formulation and its parameters can be found in the supplemental section.

**Table 1:** Parameter definitions and boundaries: This table describes the physiological interpretations of the fifteen parameters used in this model (11, 15). The range column indicates the upper and lower bounds within which an error minimizing function may establish an optimal value with respect to a concrete set of patient data. The * in place of upper and lower bounds of *A*_0_ is because the range of *A*_0_ is patient specific and is set to the patient’s maximum recorded androgen data ±10.

| Parameter | Description | Range | Unit |
| --- | --- | --- | --- |
| $M$ | max proliferation Rate | 0.001 – 0.09 | $[\text{day}]^{-1}$ |
| $q_1$ | minimum cell quota for $x_1$ to proliferate | 0.41 – 1.73 | $[\text{nmol}][\text{day}]^{-1}$ |
| $q_2$ | minimum cell quota for $x_2$ to proliferate | 0.01 – 0.41 | $[\text{nmole}][\text{day}]^{-1}$ |
| $d$ | density death rate | 0.001 – 0.30 | $[\text{L}]^{-1}[\text{day}]^{-1}$ |
| $c$ | maximum mutation rate | 0.00015 – 0.00015 | $[\text{day}]^{-1}$ |
| $K$ | half-saturation constant for mutation | 1 – 1 | $[\text{nmole}][\text{day}]^{-1}$ |
| $\gamma_1$ | androgen production by testes | 0.008 – 0.8 | $[\text{nmol}][\text{day}]^{-1}$ |
| $\gamma_2$ | androgen production rate by adrenal gland | 0.005 – 0.005 | $[\text{nmol}][\text{day}]^{-1}$ |
| $A_0$ | homeostasis serum androgen level | * | $[\text{nmol}]$ |
| $\Delta$ | androgen degradation rate | 0.03 – 0.15 | $[\text{day}]^{-1}$ |
| $b$ | baseline PSA production rate | 0.0001 – 0.1 | $[\mu\text{g}][\text{nmol}]^{-1}[\text{day}]^{-1}$ |
| $\sigma_1$ | maximum PSA production rate by $x_1$ | 0.001 – 1 | $[\mu\text{g}][\text{nmol}]^{-1}[\text{L}]^{-1}[\text{day}]^{-1}$ |
| $\sigma_2$ | maximum PSA production rate by $x_2$ | 0.001 – 1 | $[\mu\text{g}][\text{nmol}]^{-1}[\text{L}]^{-1}[\text{day}]^{-1}$ |
| $\epsilon$ | PSA clearance rate | 0.0001 – 0.1 | $[\text{day}]^{-1}$ |

We fit our model to patient data from a clinical study of IAS at the Vancouver Prostate Centre (7). Data contains longitudinal measurements of PSA and androgen for 71 patients during IAS. The information on the ultimate result of each patient’s treatment is also recorded.

We use MATLAB 2021a to perform our simulation and analysis. In particular, we use MATLAB function *fmincon* to fit model to patient data. To limit potential issues of parameter identifiability, we only estimate five key parameters (12). We fit the model to data on each treatment cycle’s on- and off-period to estimate the values of these parameters. The remaining parameters are fixed to values determined by a test-run performed over a short segment of data. Additionally, we apply more weight to the discrepancy between the model simulation and PSA data as compared to androgen data (85% to 15%, respectively). Weighted error approaches have been shown to improve overall model fitting (11). The supplemental section contains additional details regarding the data used, the method of calculating error, and other considerations of the model fitting.

We present two potential biomarkers that may be used to predict the development of resistance to ADT. Our first predictive proposed indicator is the ratio between initial and final (most recent) values of *q*_2_. Selective pressure during each treatment cycle causes the resistant subclones to become less dependent on environmental androgen through a variety of different mechanisms (49). Therefore, we expect the value of *q*_2_ to decrease over sequential estimates. We aim to determine, using clinical data fitted to the model, if one can define a threshold that is correlated with an increased probability of treatment failure.

Treatment resistance can also be recognized in the data by the divergence between androgen and PSA dynamics. Initially, when a patient begins androgen suppression therapy, the PSA and androgen behavior are qualitatively similar during on- and off-treatment. However, as the neoplasm becomes more castration resistant, more cells survive cycles of androgen deprivation to secrete PSA regardless of treatment. The second proposed biomarker is built on this observation, which takes the ratio of serum measurements of androgen and PSA to be an indicator of treatment failure. In essence, the second proposed biomarker is a model-free form of the first biomarker.

In order to evaluate the predictive potential of both proposed biomarkers, we first sorted patients into two groups: success or failure. We defined failures as discontinuations due to resistance or death from prostate cancer; we classified all other outcomes as successes. Next, we sought to identify a correlation between the biomarker values and treatment success or failure. Distinct thresholds were established for each ratio that, when reached, signify treatment failure in the next cycle.

We present here two sets of thresholds calculated with two different methods. The first method determines a threshold that maximizes the accuracy of our predictions based on the Vancouver dataset, which is denoted as the Max threshold. The second method uses MATLAB’s support vector machine function *fitcsvm* to generate a threshold that, while less accurate than the Max threshold when used with the current dataset, is potentially more accurate across a larger cohort of patient. The threshold calculated by the second method is referred to as the SVM threshold. We then test the robustness of these thresholds with cross-validation.

## Results

In general, the model closely captures PSA and androgen dynamics (Fig. 2). However, its fit favors PSA data over androgen data [11]. Nevertheless, our model is capable of capturing PSA and androgen dynamics sufficiently to generate accurate predictive biomarkers.

**Figure 2:**
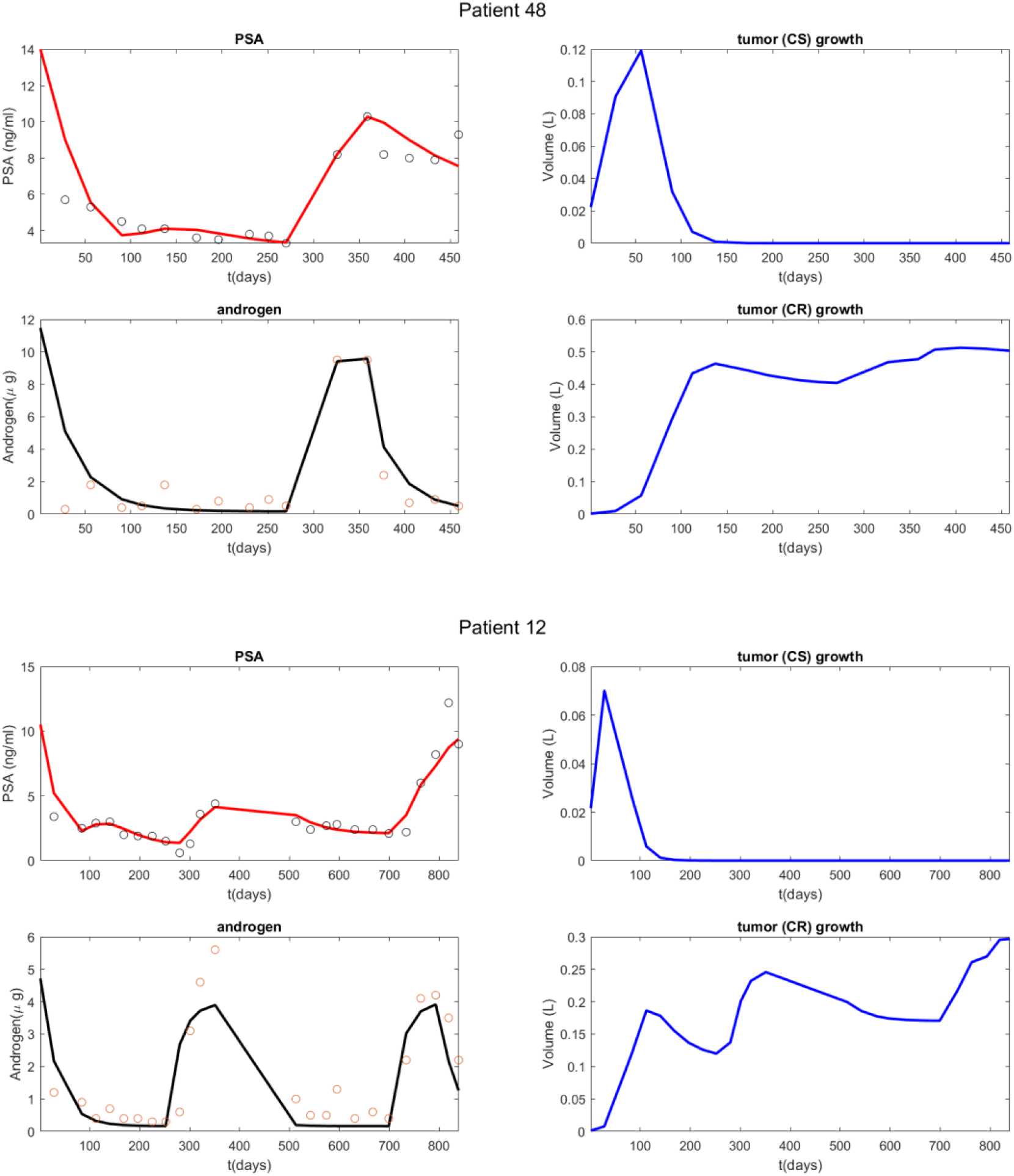
Model validation: Best-fit model solutions to the dynamics of serum androgen and PSA levels. Circles represent patient measurements, and the solid lines are solutions of model (model equation number). ‘CS’ = castration susceptible tumor cell population; ‘CR’ = castration resistant population. Panel (a) was produced by a short dataset 1.5 cycles long, and panel (b) by a dataset 2.5 cycles long.

Figure 3 summarizes the analytical result for the *q*_2_ ratio as a potential predictive biomarker. The result demonstrates that when the value of the *q*_*2*_ ratio exceeds either the SVM or Max threshold, it strongly indicates an impending treatment failure due to the development of resistance. The *q*_*2*_ Max and SVM thresholds classify the data with accuracies of 87.3% (sensitivity: 96.1%, specificity: 65%) and 81.7% (sensitivity: 98.0%, specificity: 40%), respectively.

**Figure 3:**
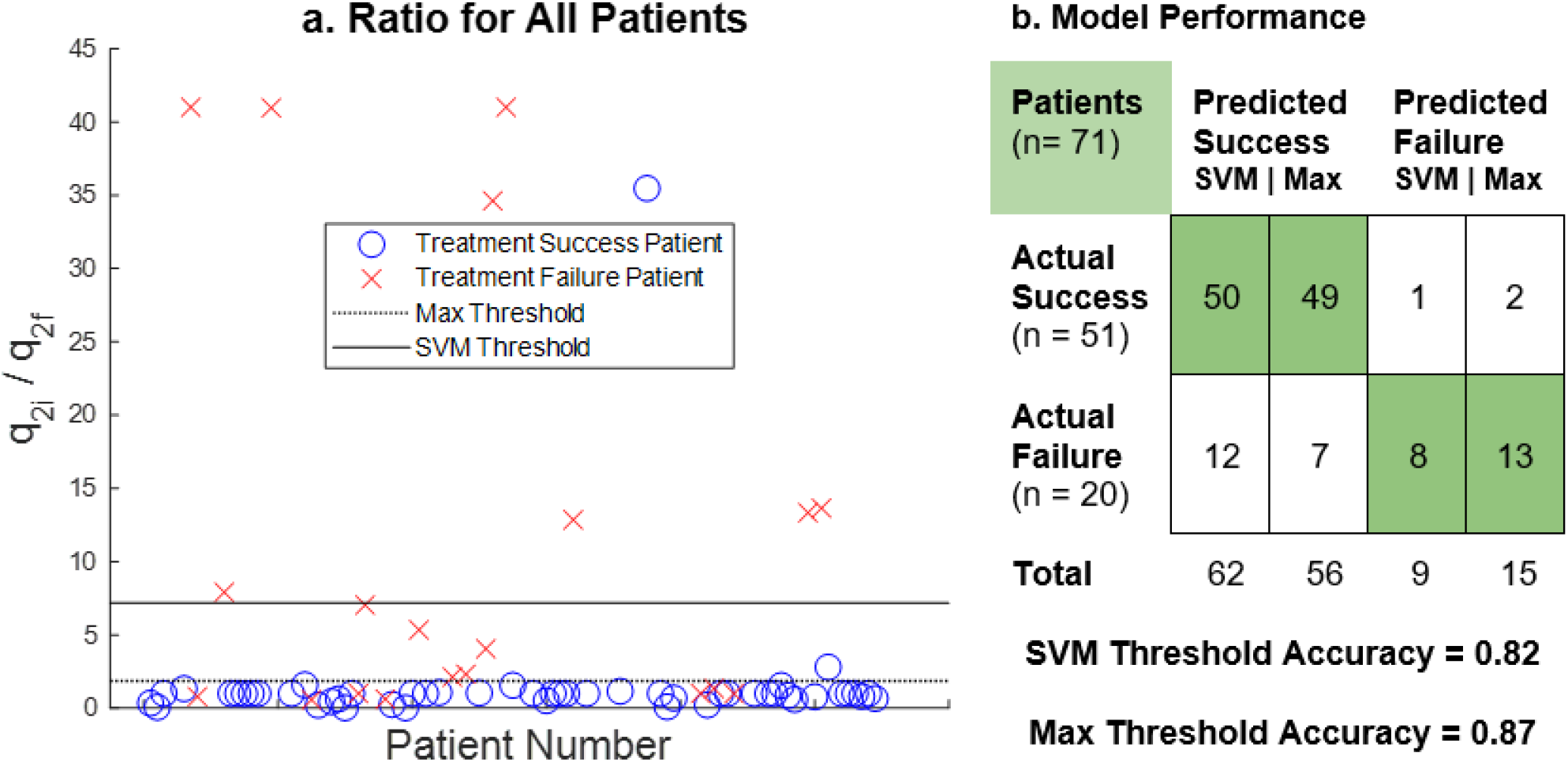
The predictive potential of the q_2_ ratio: The scatterplot (*a)* indicates the value of the *q*_*2*_ ratio for every patient in the dataset. The ratio is between the initial and final values of the *q*_*2*_ parameter calculated by the mathematical model. Max (dotted line) and SVM (solid line) threshold values are shown. The confusion matrix (*b)* compares actual patient outcomes with outcomes predicted by *q*_*2*_ ratio with respect to the thresholds.

Figure 4 shows a summary of the analytical result for the androgen to PSA ratio as a potential predictive biomarker. The initial values of the androgen/PSA ratio are highly variable across all patients. In the early stages of treatment, there is no correlation between the ratio and that treatment’s ultimate outcome, which is consistent with the selection criteria of the patients for the clinical trial (9). However, that is not the case if the androgen to PSA ratio is calculated using the mean values of the patient’s final on-treatment cycle. When the androgen to PSA ratio falls below the Max or SVM thresholds, it strongly indicates impending treatment failure due to the development of castration resistance. For the androgen to PSA ratio, the Max threshold is 0.19 and classifies patients with 88.7% accuracy (sensitivity: 90.2%, specificity: 85.0%), while the SVM threshold of 0.30 classifies patients with 84.5% accuracy (sensitivity: 82.4%, specificity: 90.0%). It is worth emphasizing that the androgen/PSA biomarker is calculated using only serum androgen and PSA data, and therefore does not require a mathematical model to estimate.

**Figure 4:**
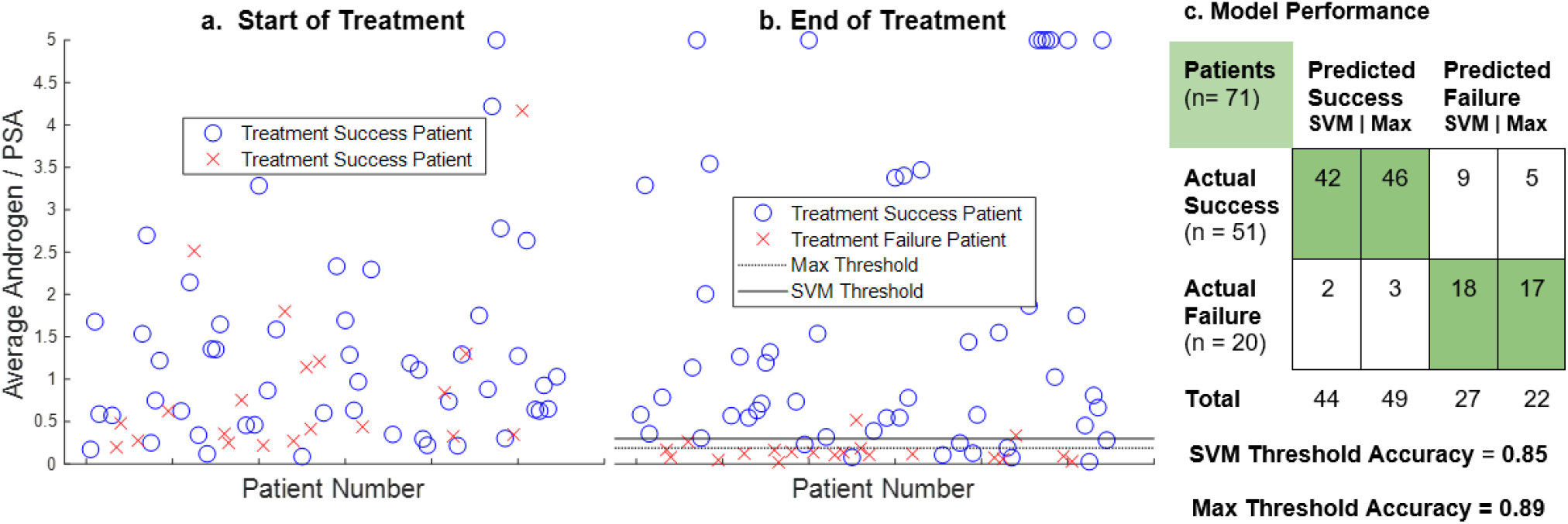
The predictive potential of the Androgen/PSA ratio: Scatterplot (*a)* shows the value of the androgen/PSA ratio for every patient when calculated using mean values of androgen and PSA from the first 200 days of treatment. Scatterplot (*a)* demonstrates that there is little correlation between the value of the ratio and treatment outcome when calculated in this manner. Scatterplot *b* shows the same ratio calculated using mean androgen and PSA values from the patient’s final on-treatment cycle, not exceeding 200 days. For the purposes of this figure all ratio values greater than five are represented as five. Scatterplot (*b)* shows two thresholds below which values of the androgen/PSA ratio indicate impending treatment failure. The confusion matrix (*c)* compares actual patient outcomes to outcomes predicted by the ratio with respect to the two thresholds.

To test robustness of all thresholds, we used a five-fold cross-validation. For the SVM thresholds, we used the built-in MATLAB cross-validation function. The predictive accuracy for the *q*_*2*_ ratio SVM threshold was 79%. The accuracy for the androgen/PSA SVM threshold was 85%. For the Max thresholds, we randomized the data, performed the five-fold cross validation, and then replicated the procedure 100 times. The mean accuracies of the *q*_*2*_ ratio SVM thresholds were 87% for the training sample, and 84% for the holdout sample. The mean accuracies for the androgen/PSA SVM thresholds were 89% for the training sample and 85% for the holdout sample.

## Discussion and Conclusion

Perhaps the most troublesome cancer characteristics, when it comes to treatment, is its ability to quickly adapt and evade initially effective treatments. Due to genomic instability, new cellular variations are continuously appearing, competing, and going extinct, leading to increasing malignancy via natural selection (4,23,24). Therefore, in many cases, it is only a matter of time before malignant tumors evolve resistance to conventional treatments. When a treatment fails, susceptible subclones have perished and been replaced by resistant clones, thus removing the possibility of continued success for first- or even second-line treatment. If clinicians could detect incipient resistance in advance, it would allow them an opportunity to change tactics that may lead to better clinical outcomes. Such an ability would likely improve long-term management and individualized treatment plans for cancer patients (17,25,32).

In this study, we propose two different tools that can be used to determine approaching castration resistance during intermittent ADT with high accuracy. Prediction tools exist to assess clinical prostate cancer, including AUC measures of PSA to diagnose clinically significant disease (18) and Gleason score, which has some power to measure prostate cancer aggressiveness and predict treatment outcome (19). However, neither PSA nor Gleason score are typically used to make real time predictions of cancer response to treatment. In contrast, our two proposed indicators rely on measurements of PSA and androgen, which are both relatively simple to obtain in real time, and have high predictive accuracies of 87 – 89%. This observation supports their potential usefulness in the clinical setting and warrants further investigation.

Both proposed biomarkers are developed from the same classical ecological theories. However, they are calculated differently. While the androgen to PSA ratio was motivated by a mathematical model, the model is not required; it relies entirely on serum data that can be collected as part of the standard monitoring routine. However, the model-free biomarker requires consistent, regular measurements of both androgen and PSA serum for calibration and prediction accuracy. In contrast, the *q*_2_ ratio may be estimated using only longitudinal PSA data and baseline serum androgen level with the aid of the mathematical model. Furthermore, since *q*_2_ reflects the degree of resistance and may be estimated independently of androgen data, it is possible to detect approaching treatment failure during the off-treatment period prior to resuming treatment. Both proposed biomarkers can be used in conjunction to improve the overall accuracy.

In our analysis, we calculate the ratios using data from the last on-treatment to show the predictive potential of our proposed biomarkers. This leaves an important question unanswered: how early can these indicators predict treatment failure with sufficiently high accuracy? We demonstrate that the proposed biomarkers do not indicate treatment failure at the start of treatment and predict treatment failure with high accuracy only at the last cycle. Furthermore, we show that there is an increasing trend in *q*_2_ ratio over each treatment interval (see supplementary figure 8s). This implies that the potential of being able to predict the treatment outcome dynamically over the course of treatment. Subsequent studies are needed to examine this aspect.

The high accuracy of these two biomarkers in our analysis supports the growing trend of implementing mathematical models in clinical studies (15,44). Furthermore, our analysis reemphasizes the importance of careful data collection during treatment. The dataset that we use here contains consistent longitudinal measurements of PSA and serum androgen for each patient over several years of treatment. However, this is not often the case in practice. For these, or any, biomarkers to have practical value, blood panels measuring serum PSA and androgen must be taken regularly and consistently to maximize the usefulness of mathematical models (12). Therefore, the development of mathematical models in clinical settings can benefit tremendously from incorporating data sets that are specifically designed and collected for the validation of the models.

## Data Availability

All data produced in the present work are available online at indicated in the data availability section of the study.

## Acknowledgements

W.M., A.W. and E.H. were supported partially by the Research Experience for Undergraduate program, (AM)^2 REU, at Arizona State University. T.P. is supported by the director fellowship at Los Alamos National Laboratory. Y.K. is partially supported by the US National Science Foundation Rules of Life program DEB -1930728 and the NIH grant 5R01GM131405-02.

The authors would also like to thank Professor Wenbo Tang and Matthew Berkes for their help during the REU program.

## Data and Code Availability Statement

All codes and results are available online at https://github.com/allison-weber/prostate_cancer_modeling_study.

## Supplemental Material

### Model Formulation

The model we present was formulated incrementally as a series of changes, modifications, and occasional failures over time. This is a summary of the steps taken to get from an established starting point to the final form of the model presented in this publication.

We began with a model published by Baez et al (1). The Baez model is a system of four differential equations that represent: the volumes of two cancer cell subpopulations, the cell quota of androgen, and the serum concentration of PSA. The model is as follows:

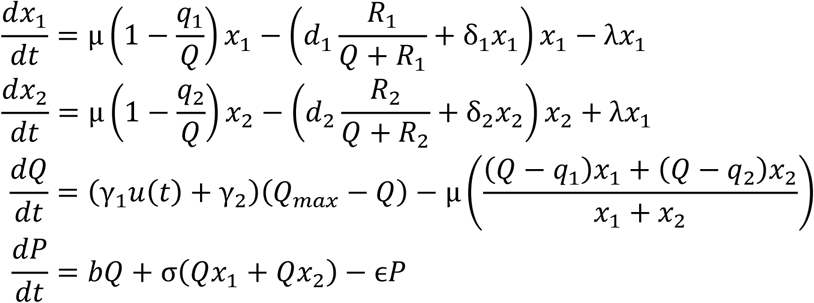

When we reproduced the Baez model, we found that we could rarely recreate the whole range of androgen and PSA data. Peaks of data caused by spikes in data values were often truncated to some lower level. Our original goal was to discover if adjustments could be made that would allow the model to better represent these peaks.

**Figure 1s:**
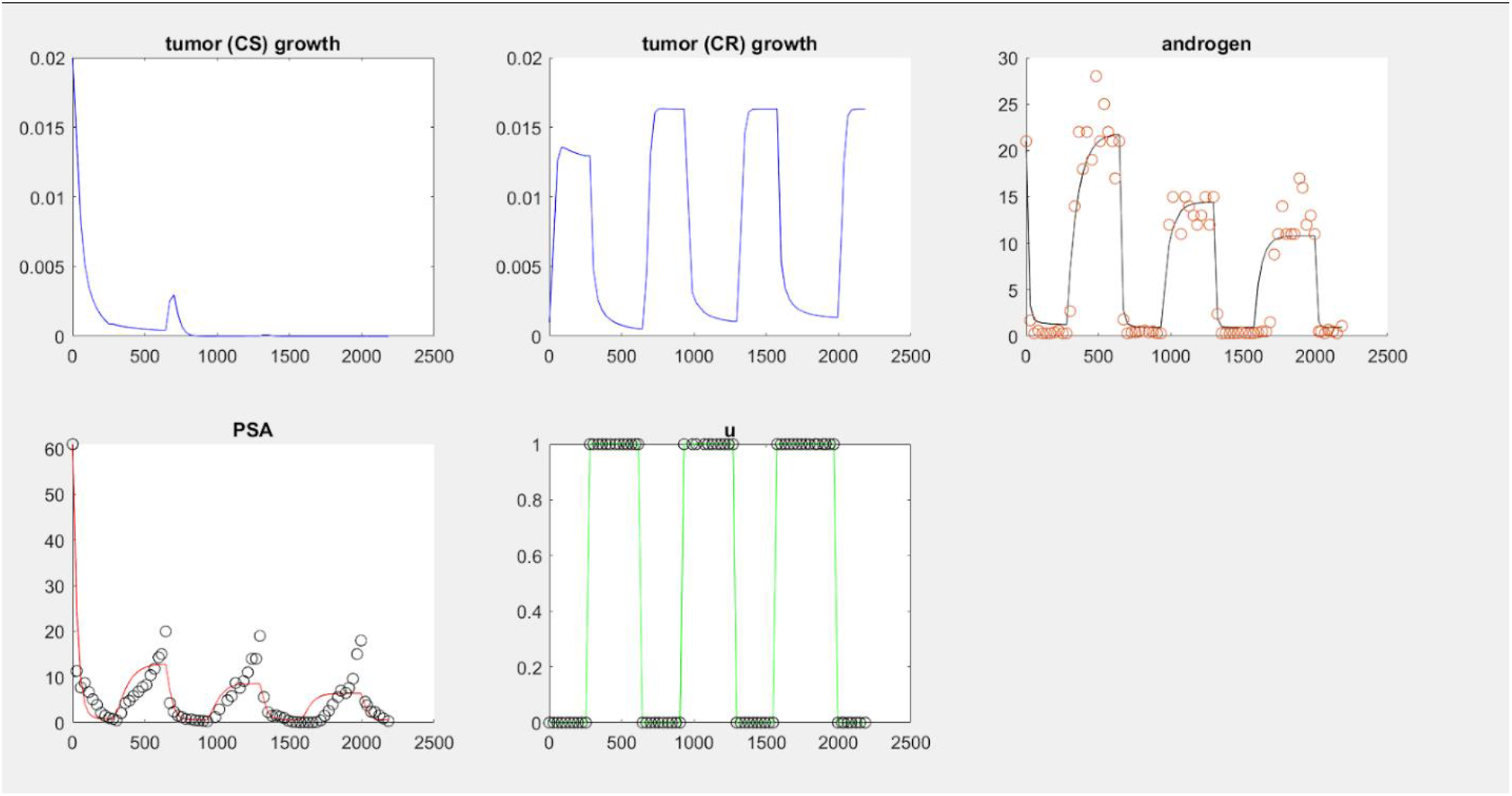
This one result of our recreation of the Baez model. The data are represented by black circles, while the simulated results are solid lines. The figure titled ‘u’ shows when the patient was on or off treatment. This purpose of this figure is to show that, in the case of both androgen and PSA, the model was not reproducing the entire range of data. The upper end of measurements, or ‘peaks’, are cut off.

To that end, our first change was to condense the two death rate parameters into just one term. When we examined the optimized parameters returned to us by the fmincon function, we noticed that there was always very little difference between the values of *d*_*1*_ and *d*_*2*_. We decided that the difference was small enough to justify merging these terms so that the model might better identify the other parameters. On the other hand, we also split the σ parameter into *σ*_1_ and *σ*_2_. We did this to allow for the possibility that both subclones were producing PSA at different rates. This is what the model looked like after making those changes.

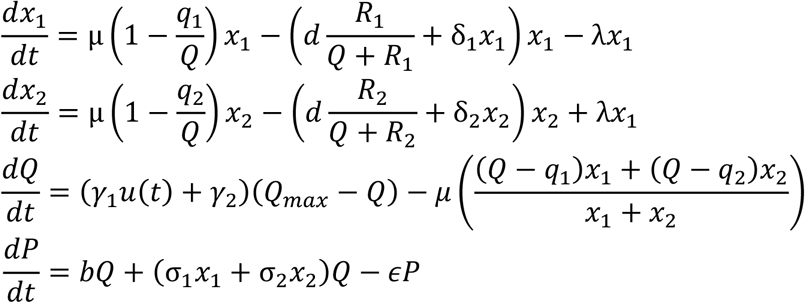

**Figure 2s:**
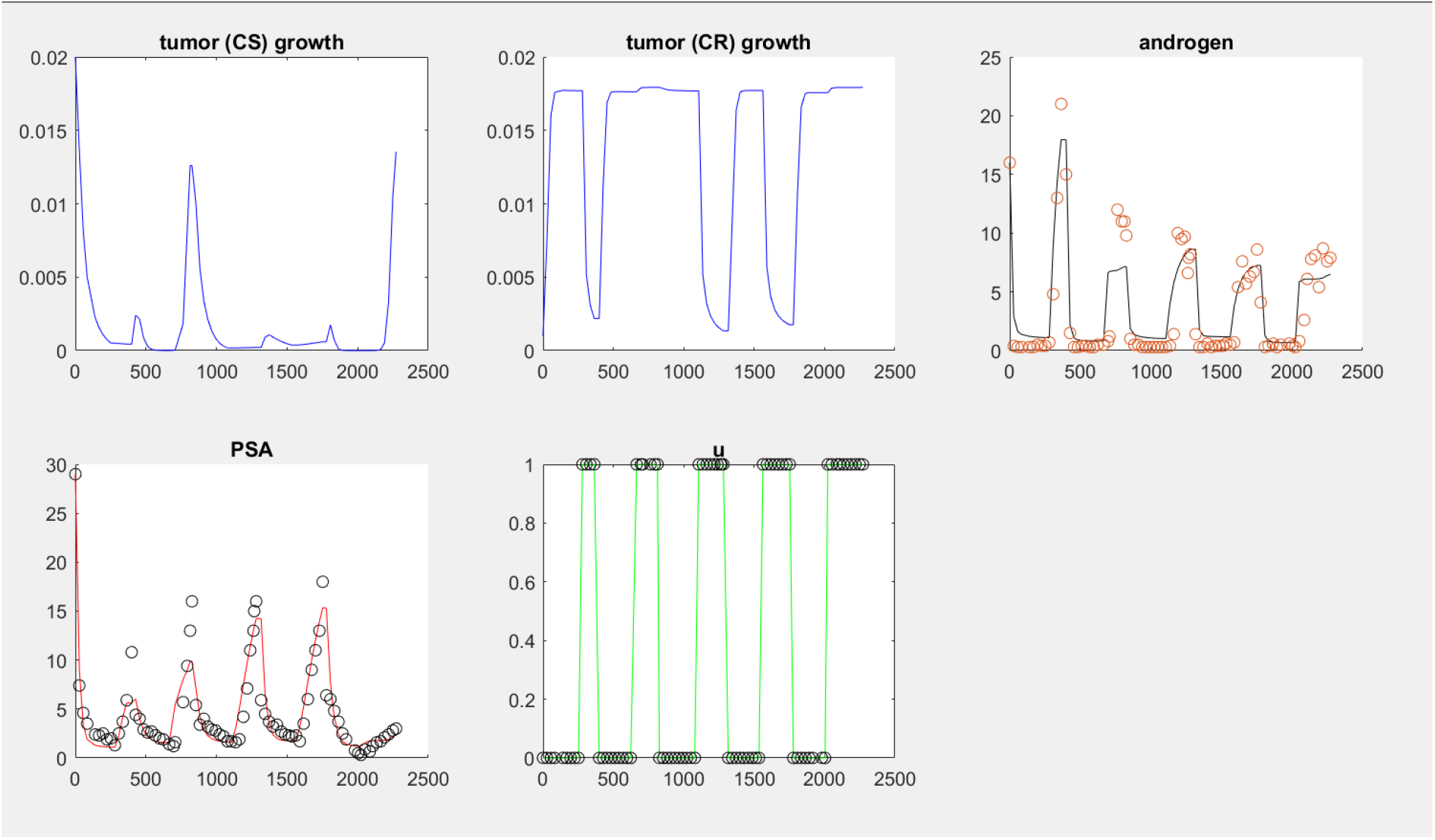
An example of the results produced by the first series of modifications to the Baez model. The two death terms, *d*_*1*_ and *d*_*2*_, were merged into one, *d*, and the σ parameter was split into σ_1_ and σ_2_. The apparent result of these two adjustments was an improved capacity for the model to represent the data.

Taken together, these two changes were found to somewhat improve the model’s ability to simulate larger measurements. An example of the results may be seen in Figure 2s. These changes were tested separately, and it was found that making the change to σ alone was not sufficient to apparently improve the result.

Next, we implemented three substantial modifications. These changes were tentative and experimental, so they were all tested separately and in combination with each other. In addition, all of these model variations were tested with and without the addition of a degradation term to the *dQ* equation.

The first modification was the complete removal of the death terms. Negative growth is already made possible by the Droop equations in the growth terms (growth is negative when *Q*< *q*_i_), and we wanted to see if this negative pressure would be sufficient to control the cell populations without the death terms.

The second modification was the removal of the *dx*_*2*_ equation. We wanted to know exactly how much the second layer of complexity contributed to the outcome. We thought it possible, since we were continually fitting the parameter *q*, that resistance might be adequately modeled by the dynamics of *q* alone. If we could remove one of the differential equations without substantially damaging the result, it would make it much easier for fmincon to identify the remaining critical parameters (see section below for more information on fitting and critical parameters).

The third modification was a substantial change to the PSA equation and involved incorporating the Droop functions from the growth terms into the PSA production terms. This is a novel modification that introduces an entirely different interpretation of what motivates PSA production and is discussed in the main body of this publication.

This is the system of differential equations that incorporates all three of these modifications, and includes the degradation term in the *dQ* equation:

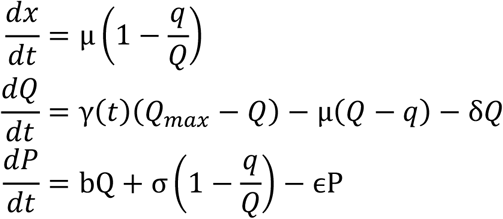

The products of this system were surprising, but still useful, and may be seen in Figure 3s. Clearly this is a failed result, but it was immediately apparent where we had made the mistake: by applying the Droop functions to the *dP* equation, we had conferred the possibility for negative growth (due to a lack of androgen) to PSA as well. Furthermore, we can see that by removing the death terms that the tumor volumes and PSA concentrations have both grown far beyond the limits of believability.

**Figure 3s:**
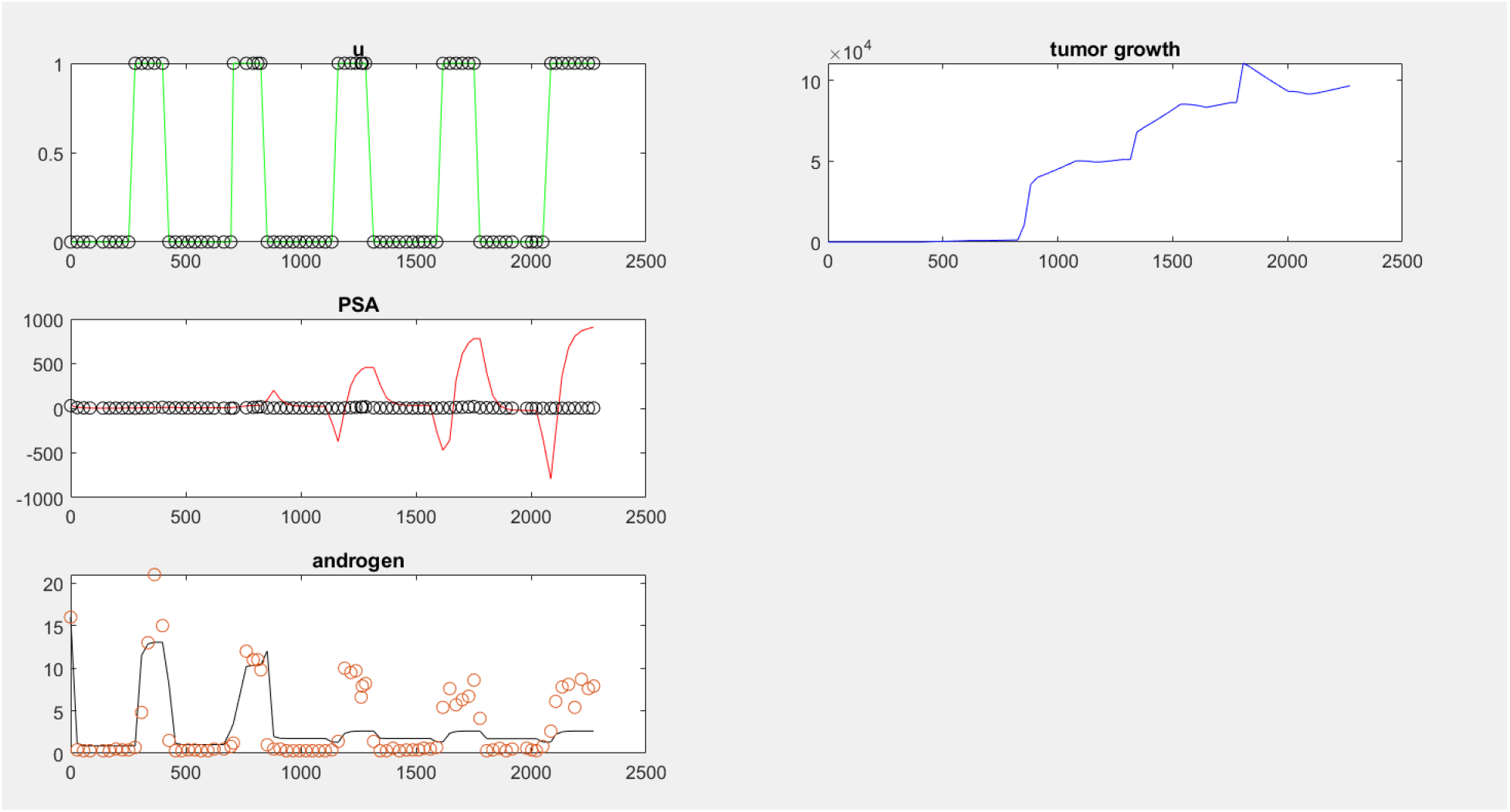
This figure shows the result of removing the death terms and applying Droop functions to PSA production. This is clearly a failed attempt. PSA oscillates between positive and negative values, which is clearly impossible. Furthermore, by the end of treatment the magnitude of the tumor volume is entirely incredulous.

To rectify this mistake, we applied maximum functions to all the related Droop terms. Negative growth needed to be eliminated from PSA production, but for consistency we applied the maximum functions to the growth terms as well. In doing so, we eliminated the built-in capacity for negative growth in the *dx* equations, where it was needed. This, in addition to the explosive, runaway growth of the cell volumes seen the previous result, required us to reintroduce death terms to the model.

We initially tested two variations on these death terms, *d*_1_*x*_1_ vs 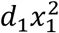, and found that the latter was necessary to keep the populations under control. When we reintroduced these death terms we used two separate death rate parameters: *d*_*1*_ and *d*_*2*_.

After experimenting with all the variations, and implementing these corrections, the best performing model was this (see also Figure 4s):

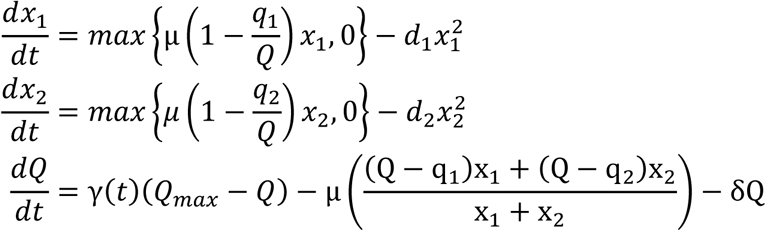

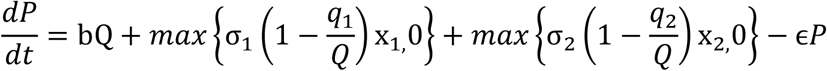

**Figure 4s:**
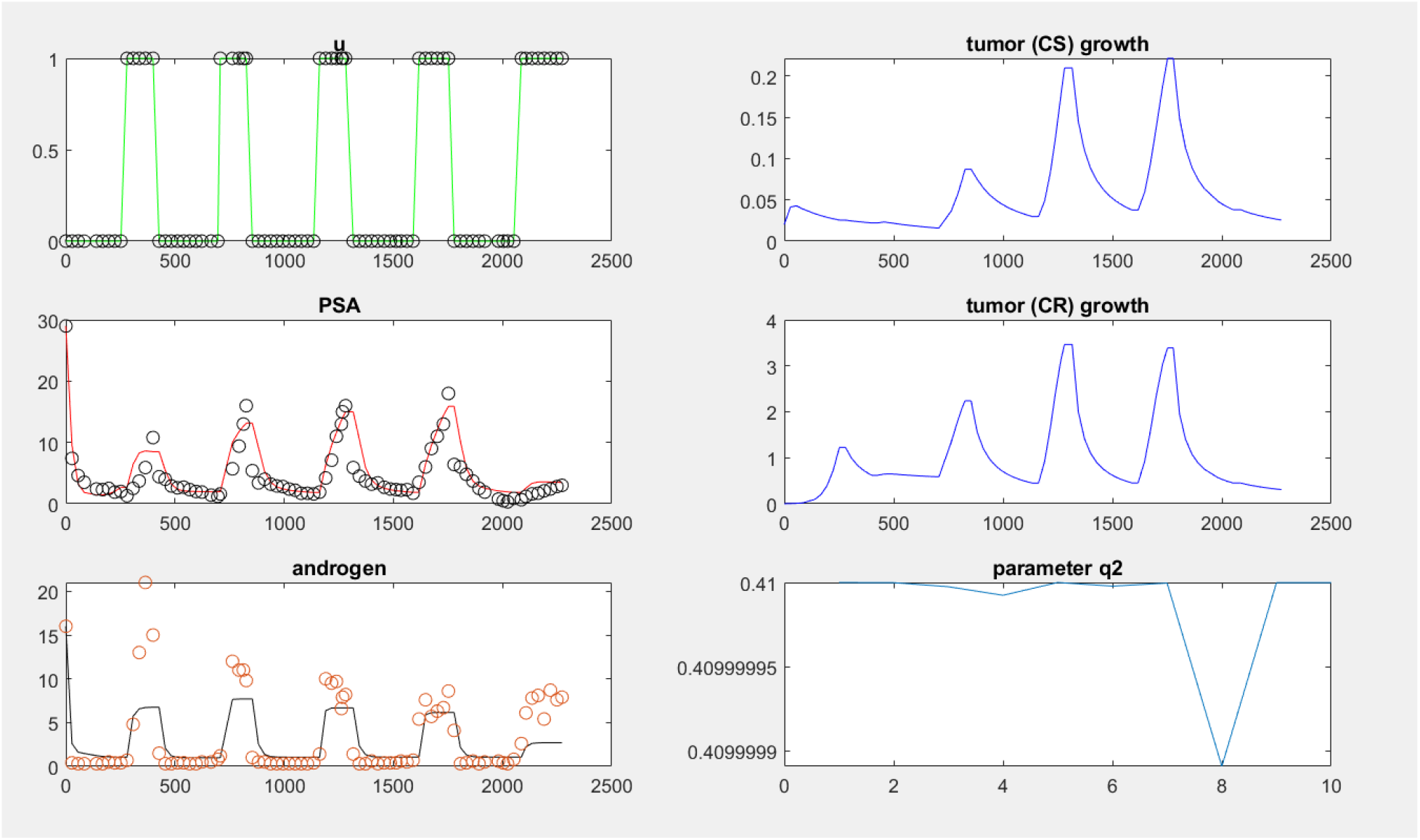
This figure is an example of the result produced by the model when the Droop functions were applied to the PSA production terms and upon reintroducing death terms to the growth equations. This result was viewed as a success, but we identified a remaining challenge in that the volume of the resistant cell colony was still unrealistically large.

Our next challenge was that the simulated tumor volumes were often impossibly large. In Figure 4s, for example, we see that the volume of the treatment-resistant subpopulation is approaching four liters. Our next task was to implement changes that might affect a reduction in those numbers.

We first tested a range of new values for the death rate parameters, with varying levels of success, before again merging them into a single parameter *d*. We also tested a range of new values for the growth parameter μ. Unable to find the bounds for *μ* that gave the desired result, we incorporated a change in how the *μ* parameter was optimized. Whereas before we programmed fmincon to optimize μ, alongside other parameters, against a short, two-cycle test segment, the program was changed such that *μ* was fitted first, before any other parameters, against an even shorter, half-cycle segment. The resulting value of *μ* was then fixed before any other parameters were optimized. This was found to successfully reduce the upper bounds of our tumor volumes without substantially changing the underlying dynamics.

**Figure 5s:**
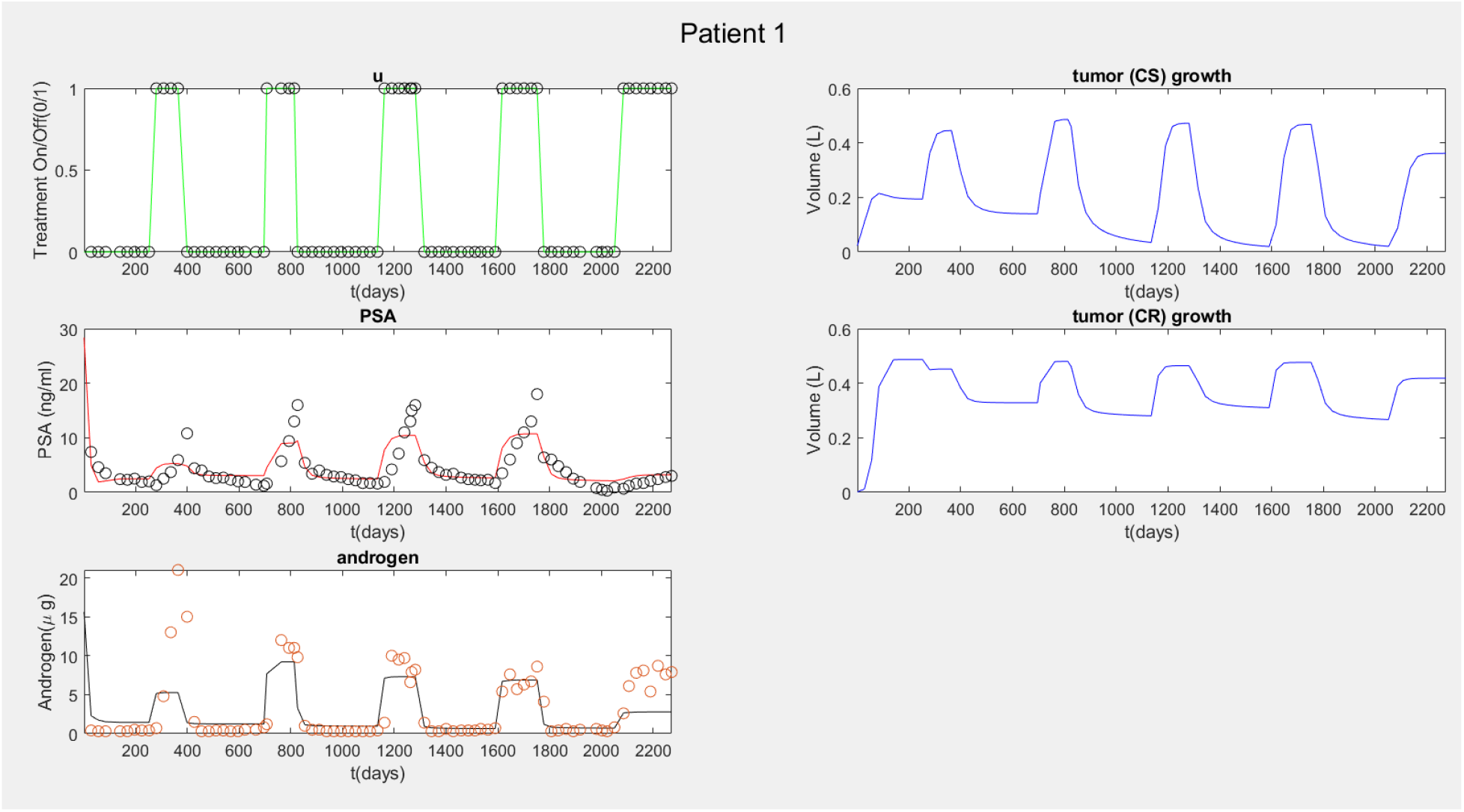
Changing the method of optimizing the single parameter *μ*resulted in a more realistic range of resistant tumor volume. Notable is that the CS and CR populations co-exist throughout the course of treatment, with one never overtaking the other.

Feeling that the volumes of *x*_*1*_ and *x*_*2*_ were now under control, we moved on to something new. We noticed that the resistant and susceptible tumor populations, *x*_*1*_ and *x*_*2*_, often coexisted together throughout the course of the treatment. This can be seen in Figure 5s where, although both populations grow and shrink according to treatment status, one is never seen to outcompete the other. This did not conform to our expectations, which were for the resistant subpopulation to become dominant as treatment is applied repeatedly over time. To address this, we decided to modify the death terms once again, this time to introduce an element of interspecific competition between these two subpopulations.

These are the first two differential equations after the introduction of interspecific competition:

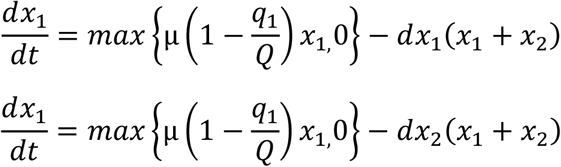

Implementing this change had some of the desired effect on the dynamics of the susceptible population. Figure 6s shows that, upon the implementation of these new death terms, the susceptible population was indeed seen to go extinct. This was, however, only a partial success. We would have preferred to see less oscillation in the resistant population, and for the susceptible population to fail over a longer period. Ideally, we want to see that PSA dynamics are driven by oscillations in the susceptible population, and that extinction of the susceptible population coincided with a deviation between androgen and PSA behavior. This is open challenge that we may address in the future.

At the time, however, the primary problem was that by introducing interspecific competition we had significantly damaged the model’s ability to reproduce androgen data. Therefore, we undertook one final modification in the hopes of improving upon that shortcoming: the introduction of a fifth differential equation.

**Figure 6s:**
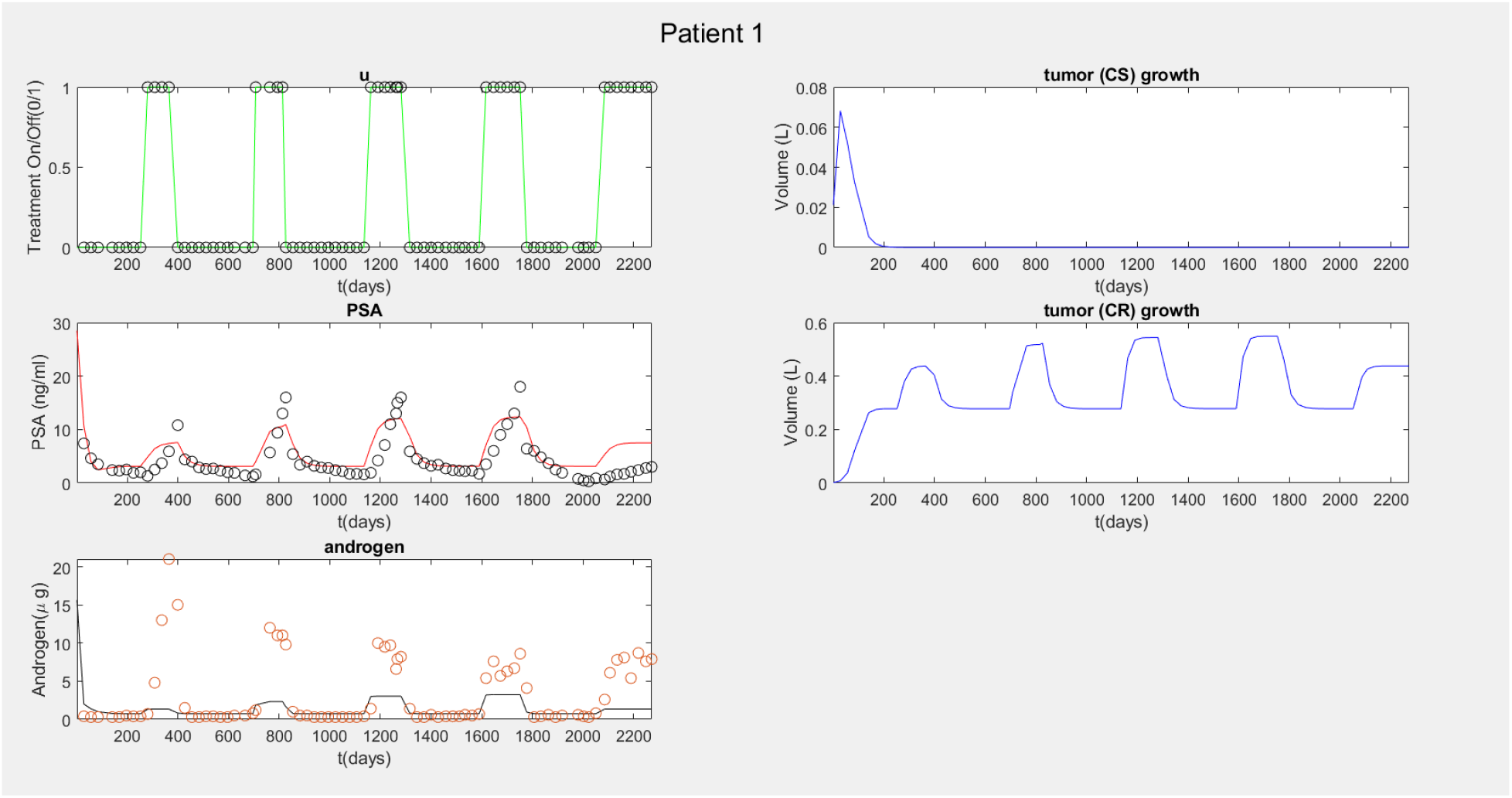
This figure shows the result of introducing interspecific competition. The susceptible subpopulation is now seen to die off, as expected. This result is only a partial success. The desired outcome was achieved, but likely too early, and too abruptly. We would also have preferred to see less oscillation in the resistant subpopulation. Finally, the ability of the model to reproduce androgen values has noticeably worsened.

The purpose of introducing a fifth differential equation was to compartmentalize the serum androgen, for which we had measurements, from the cell quota of androgen that motivated tumor growth and PSA production. We tested two different forms of this fifth differential equation. The first was published by Phan et al, and the second by Reckell et al (2,3). We found that, in the context of this model, the second equation produced better results.

Androgen equation (Phan et al):

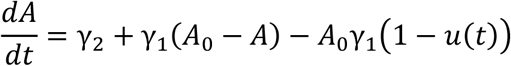

Androgen equation (Reckell et al):

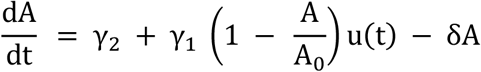

Implementing this fifth differential equation gives the final form of the model presented in the main body of the publication. Using this model, we were able to accomplish some of the goals we had when we began, but there remain opportunities for improvement. Upon introducing the fifth differential equation, we saw that the volumes of the resistant subpopulations again started to approach unrealistically large values. Furthermore, the model still struggles to reproduce androgen data for many of the patient datasets we tested. This is, to an extent, an expected consequence of how we weigh error (80% in favor of PSA), but we still believe that improvement is possible.

**Figure 7s:**
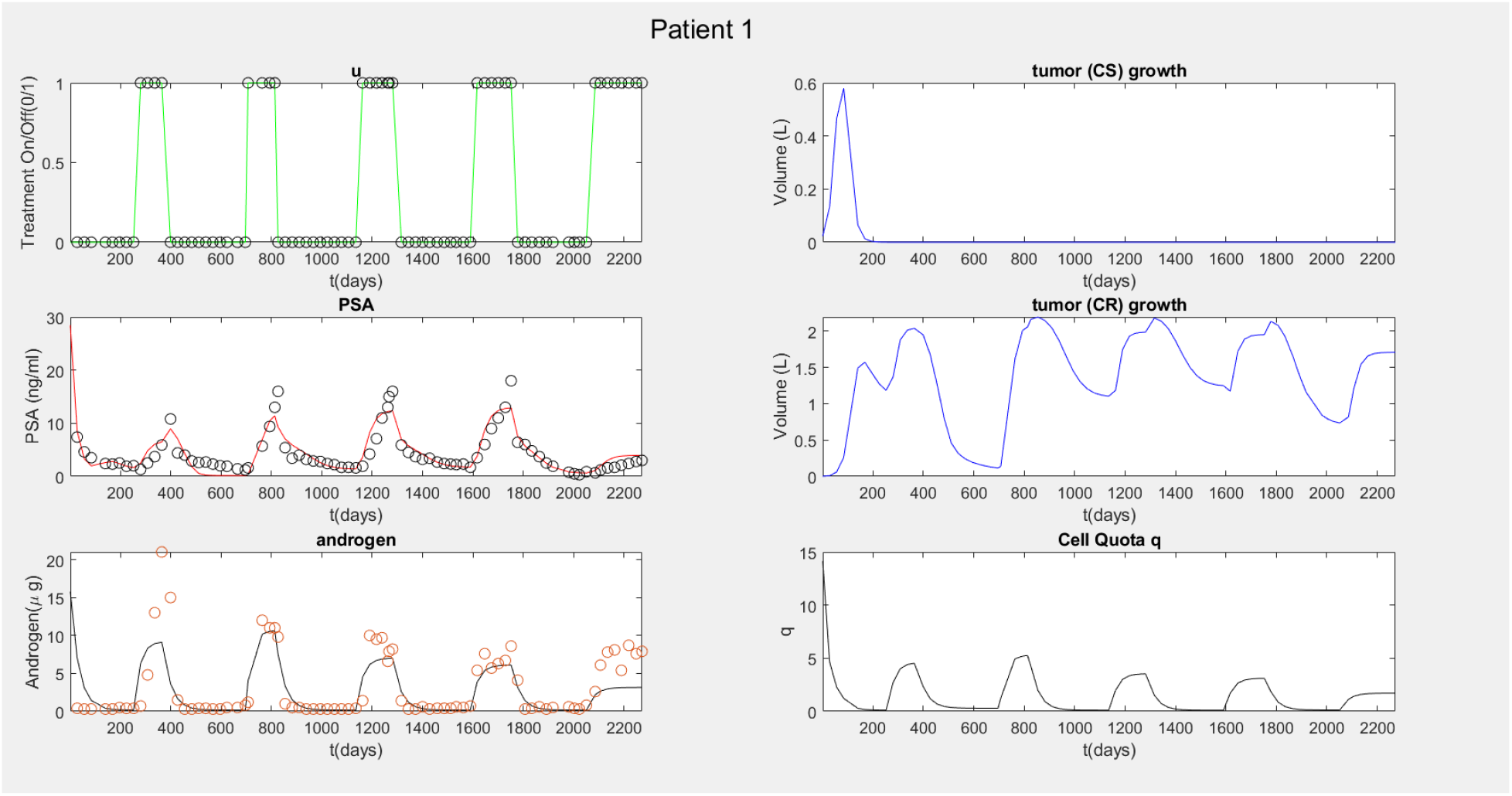
One result of compartmentalizing serum androgen from cell quota of androgen. The model’s ability to recreate androgen has been improved, but the volumes of the resistant subpopulation have again grown to levels that may yet be too large. This is the final form of the model presented in this publication.

### Objective Function

Essential for the operation of the Matlab’s fmincon function is the definition of the objective function, used to calculate the discrepancy between the model output and the actual data. We used the following objective function:

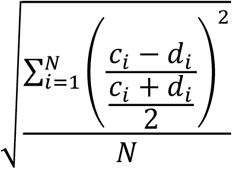

We chose to use this rather than something like sum of squared errors because we did not want the function to disproportionally weigh the largest errors. This reduced the power of outliers to disproportionally influence the optimized parameters. For the purposes of this investigation, PSA error was weighed far more heavily than androgen error (80% vs 20%).

### Parameters and Fitting

The parameter values and ranges used in this model follow directly from work done by Phan et al (2).

Three of the fifteen parameters used in this model, *c, K*, and *γ*_2,_ are permanently fixed at some static value. We do this following the sensitivity analysis done by Phan et al, which led to them permanently fixing five of their parameters: *c, K, δ*_*1*_, *δ*_*2*_, and *γ*_2_ (Note: Where they use *δ* for the density-dependent death rate, we use *d*) (2).

The mutation rate parameters *c* and *K* govern the rate that members of the susceptible population dynamically respond to treatment, and therefore move from the treatment susceptible to the treatment resistant population. The sensitivity analysis done by Phan et al demonstrated that these parameters were particularly insensitive, and that fixing these parameters was therefore a reliable way to improve the identifiability of the more critical parameters. A basis for the range of *c* was first published by Ideta et al (4). Following that, subsequent models, including this model, have used a fixed value of 0.00015 for *c*, and of 1 for *K* (1,2,5).

The next parameter *γ*_2_ stands for the rate of secondary (adrenal) androgen production. This parameter is so insensitive that Phan et al set it to zero in their work (2). We did not set it to zero, but instead fixed it such that it would be a small percentage of *γ*_1_, the primary androgen production parameter.

When it comes to the density death rate parameter *d* we deviate from the values used in earlier publications. The changes made to this model, and in particular the introduction of interspecific competition, require entirely new parameters. We therefore had no basis for setting a fixed value for *d* and allowed the parameter to be fitted by fmincon.

The twelve remaining parameters, including *d*, are all fitted in some way by fmincon. These parameters may be sorted into three different groups: four critical parameters, μin a category by itself, and seven partially fitted parameters. The four parameters most sensitive and most important to our investigation are re-optimized for every half-cycle of treatment. In other words, every time the data indicates a switch in treatment status fmincon calculates a new best-fit value for that segment of treatment. This allows us to study the dynamics of these parameters over the whole course of treatment. μ is only fitted against the first half-cycle of treatment. The program runs for one half-cycle, discovers a set of best-fit parameters for that single half-cycle, and then fixes μ at that value. The seven remaining parameters are fitted against a two-cycle test segment, and then fixed at the resultant optimal value. In order, first the program runs the single half-cycle to find the fixed value of μ, then it runs for two whole cycles to find the fixed values of the seven partially-fitted parameters, and then the program runs the whole dataset, fitting and re-fitting only the four critical parameters.

The four sensitive, critical parameters are: *q*_*2*_, representing the minimum amount of androgen required by the resistant cell population to survive and proliferate; *γ*_1_, the primary (testicular) rate of androgen production; *A*_*0*_, the maximum serum level of androgen; and σ_2_, the rate of PSA production by the resistant cell population.

We cannot overstate the importance of the parameters *q*_*2*_ and σ_2_ to this investigation. It is *q*_*2*_ that is most responsible for simulating the development of resistance, and it forms a critical component of one of our proposed predictive indicators. Likewise, σ_2_ is directly related to resistance, because it is a key driver of the divergence between androgen and PSA seen in patients as they become resistant to treatment. It is due to their direct effect on the modeling of resistance that these two parameters are considered critically important, and therefore re-fitted against every half-cycle of treatment.

We use the same upper and lower bounds for *q*_*1*_ and *q*_*2*_ as those published by Phan et al (2). They discovered work wherein was published the lowest and highest recorded serum androgen levels caused by hormonal therapy (0.41 nMol/L and 1.73 nMol/L) (6). We invariably see that androgen suppression therapy initially succeeds at reducing the volume of a tumor, so we may safely infer that the minimum cell quota for the susceptible population, *q*_*1*_, lies somewhere in that range. Furthermore, we assume that *q*_*2*_ is always less than *q*_*1*_, and therefore set the upper bound of *q*_*2*_ at the lower bound of *q*_*1*_. There is no data to support the range of *q*_*2*_, and so the lower bound is an estimate.

The maximum serum androgen, *A*_*0*_, is one of the critical parameters because it is both patient-specific and especially sensitive. The data demonstrates that although values are typically below 27nm/mole, there is still considerable variation in each patient’s maximum serum androgen measurement (7). For the purposes of this investigation, each patient’s maximum measurement was identified and the upper and lower bounds of *A*_*0*_, for that patient, were set to be ±10nMol from that maximum.

The parameter *γ*_1_ has a singular impact on the shape and behavior of simulated androgen and PSA levels. Unfortunately, there is no known clinical value for the rate of testicular androgen production. This parameter is considered one our critical parameters both because of its sensitivity and because we have no basis from which to estimate its value. In their work, Phan et al started from a range published by Ideta et al, and then reduced that range until it suited their investigation (2,4). We follow their lead and use the same upper and lower bounds.

Regarding the remaining parameters: the sequence of modifications made in the formulation of our model have left us unable to use prior, published values for the density death rate parameter *d*. Therefore, we tested a range of estimated bounds, and selected the best performers. The bounds of *q*_*1*_ are identical to the bounds of *q*_*2*_, which have already been discussed. Likewise, the bounds for σ_1_ are identical to the bounds for σ_2_. Like *d*, we tested the upper and lower bounds of *μ* by testing a range of estimated values. In the end, however, we determined that the best performing bounds were the same as those published by Phan et al (2). Those bounds for μwere derived from work done by Berges et al, wherein they measured the proliferation rates of prostate cancer cell populations in both androgen-rich and androgen-poor environments (8). The work to establish the bounds of the remaining parameters: ϵ, *b, m*, and *δ*, was done by Baez et al, and Portz et al (1,5).

### Trend of *q*_2_

We examine the evolution of the cell quota parameter for the resistant cancer population, *q*_2_. In theory, *q*_2_should decrease with each treatment, making the ratio *q*_1_/*q*_*n*_ an increasing sequence where n is the current treatment cycle. We fit a linear model to this ratio for all patients.

**Figure 8s:**
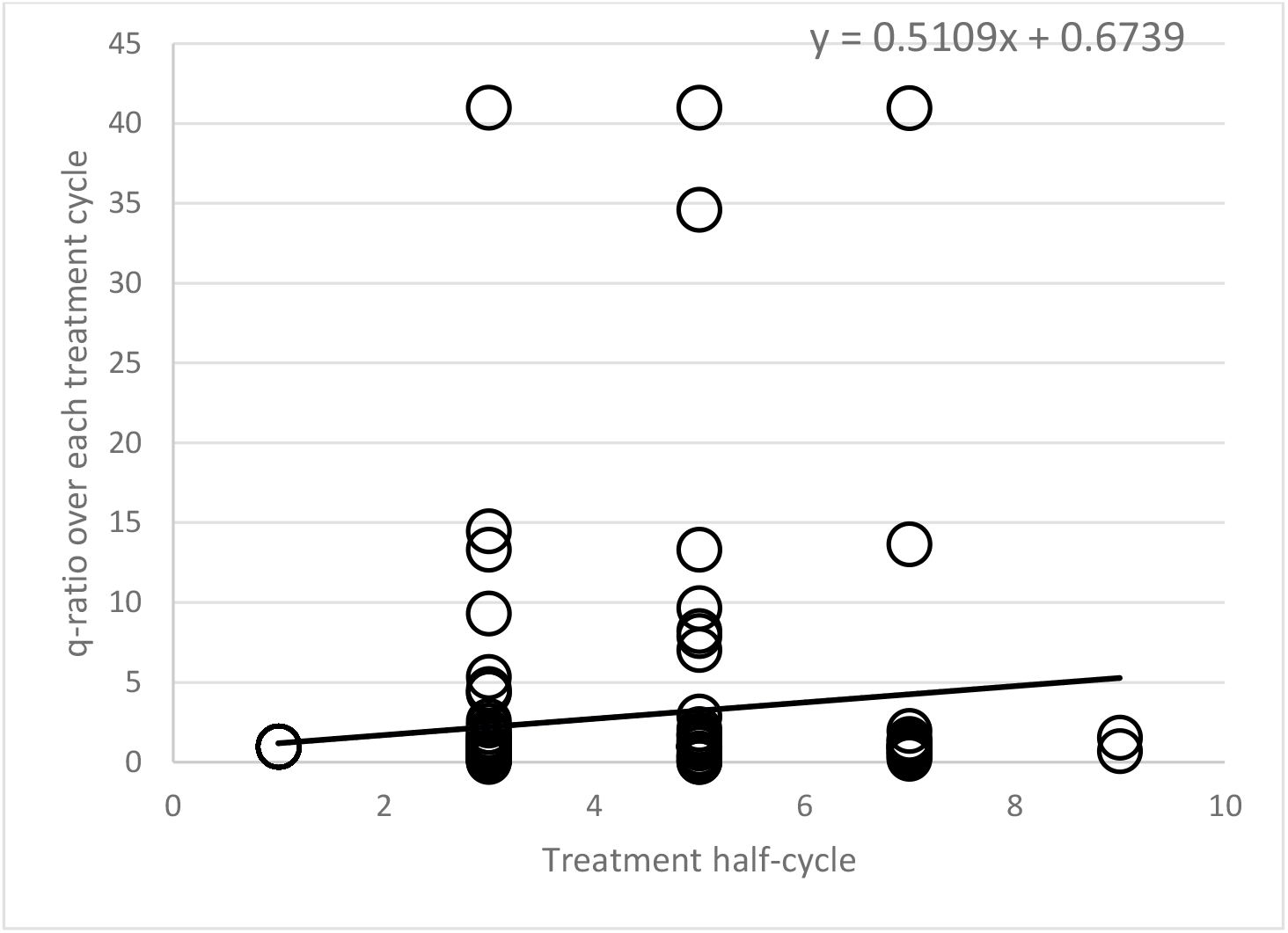
increasing trend of *q*_2_over each treatment cycle.

